# Continuous Glucose Monitoring Reveals Glycemic Patterns Associated with End-Organ Alterations in Early Dysglycemia

**DOI:** 10.64898/2026.08.14.26360480

**Authors:** Bill Chen, Anastasia-Stefania Alexopoulos, Wai Tak Lau, Kaveri A. Thakoor, Cecilia S. Lee, Ahmed A. Metwally, Jessilyn P. Dunn

**Affiliations:** Department of Biomedical Engineering, Duke University, Durham, NC; Department of Medicine, Division of Endocrinology, Metabolism and Nutrition, Duke University, Durham, NC; Department of Computer Science, Columbia University, New York, NY; Department of Biomedical Engineering, Columbia University, New York, NY; Department of Ophthalmology, Columbia University, New York, NY; John F. Hardesty MS Department of Ophthalmology and Visual Sciences, Washington University in St. Louis, St. Louis, MO; Google Research, Mountain View, CA; Systems and Biomedical Engineering Department, Cairo University, Giza, Egypt; Department of Biostatistics and Bioinformatics, Duke University, Durham, NC

## Abstract

**Objective:** To determine whether continuous glucose monitoring (CGM) identifies clinically relevant glycemic heterogeneity and subclinical end-organ alterations in adults without diabetes.

**Research Design and Methods:** We analyzed 1,017 AI-READI Year 3 participants without diabetes (558 with normoglycemia and 459 with prediabetes by A1C). Fifty-two metrics from 10-day blinded CGM were reduced to nonredundant glycemic axes. Partial Spearman correlations between representative CGM metrics and clinical measures across 13 domains were adjusted for age, sex, and BMI and controlled for false discovery rate. CGM-derived subphenotypes were identified using unsupervised UMAP-HDBSCAN-based clustering.

**Results:** Among 462 glycemic-clinical associations tested, 99 (21.4%) remained significant after false discovery rate correction. Hyperglycemia-related metrics, including mean glucose, time above range, and time in tight range, showed more associations than variability metrics. The strongest signals involved cardiometabolic, cardiovascular, and cognitive measures. Greater hyperglycemia and glucose excursions were associated with lower language performance, slower processing speed, and lower cognitive efficiency (ρ ≈ −0.10 to −0.14; all P < 0.01). Clustering identified four reproducible glycemic subphenotypes: Healthy, Mild Hyperglycemia, High Variability, and Hyperglycemia. CGM phenotypes reclassified A1C-defined groups: 58.1% of participants with normoglycemia fell into dysglycemic phenotypes, whereas 18.8% of participants with prediabetes fell into more favorable phenotypes. The Hyperglycemia phenotype had the most adverse cardiometabolic profile and lower cognitive performance.

**Conclusions:** In adults without diabetes, CGM revealed glycemic patterns associated with distinct subclinical alterations. CGM-based phenotyping may complement A1C for characterizing early dysglycemia and selecting individuals for longitudinal risk-stratification studies.

## Introduction

Diabetes affects an estimated 589 million adults worldwide, and more than 600 million individuals meet criteria for prediabetes (1,2). Although prediabetes is often conceptualized as a transitional state preceding overt type 2 diabetes, epidemiologic evidence indicates that individuals within this range already exhibit elevated risk for cardiovascular disease (3), chronic kidney disease (4), retinopathy (5), and subtle cognitive impairment (6). Continuous Glucose Monitors (CGM) have become a cornerstone of diabetes management and are increasingly integrated into clinical trials and routine care (7). Recent approval in 2024 of over-the-counter CGMs by the U.S. Food and Drug Administration further expanded their accessibility and applications beyond diagnosed diabetes (8). While A1C remains a cornerstone of guideline-based diagnosis of prediabetes/diabetes, it has several limitations, including inability to detect glycemic variation and day-to-day trends. CGM-derived metrics can address these limitations by capturing dysglycemia axes (i.e., glucose variability and excursion) that are not fully reflected by the average glycemia captured by A1C (9).

Recent work has identified the presence of early dysglycemia, defined as dysregulated glucose across the A1C-defined normoglycemia and prediabetes ranges, suggesting metabolic derangements that precede conventional diagnostic thresholds (10,11). Furthermore, studies in populations with early dysglycemia have demonstrated that glycemic axes are differentially associated with end-organ alterations and clinical outcomes (12,13). For instance, in the CGMap study, central tendencies of glucose levels were more strongly correlated with anthropometrics and body composition, whereas glycemic variability was more closely correlated with liver function (13). These findings suggest that early dysglycemia represents a heterogeneous and clinically meaningful state with physiological consequences.

Given the heterogeneity observed across glycemic axes, an emerging approach is to characterize individuals according to glycemic subphenotypes, a data-driven framework that translates glycemic heterogeneity into stratified groups for refined risk assessment and targeted interventions. In individuals with type 2 diabetes, such approaches have identified clinically distinct subphenotypes (14). Subphenotyping in largely normoglycemic populations has similarly revealed distinct patterns of dysglycemia, underscoring that conceptualizing early dysglycemia as a continuum rather than a threshold-defined state may enable more precise and proactive care strategies (10,12,15).

Despite these advances, critical knowledge gaps remain in understanding the subclinical end-organ consequences of dysglycemic stress. Prior research linking CGM patterns to end-organ alteration has focused on a relatively narrow set of clinical measures, leaving associations with other physiological systems such as cardiac structure, retinal microvasculature, and cognitive function underexplored. Furthermore, existing CGM characterization studies have primarily included individuals with normoglycemia and racially homogeneous populations, leaving potentially key demographic covariates uninvestigated in the context of early dysglycemia. To address these gaps, this study utilizes the Artificial Intelligence Ready and Equitable Atlas for Diabetes Insights (AI-READI) Year 3 (N_normoglycemia_=558, N_prediabetes_=459) dataset (16,17) to evaluate CGM-derived glycemic axes against clinical measures of end-organ alteration in a glycemically and demographically heterogeneous cohort. Accordingly, our objective is to determine whether CGM can identify clinically relevant glycemic heterogeneity and corresponding subclinical end-organ alterations in adults without diabetes.

## Methods

### Data Source

The data analyzed in this study were obtained from AI-READI Y3 (16,17); a diverse, national cohort with multimodal collection of metabolic health data. Detailed descriptions of the study design, recruitment procedures, inclusion and exclusion criteria, and measurement protocols have been published previously (18). Participants wore a blinded Dexcom G6 CGM device for 10 days. The AI-READI protocol includes standardized clinical assessments spanning cardiometabolic, renal, hepatic, hematologic, cognitive, and vision, as described in the primary study report (Fig 1A). For the present analysis, we excluded participants with missing CGM data, missing clinical assessments, A1C ≥ 6.5%, or a self-reported prior diagnosis of type 2 diabetes. Participants were classified according to American Diabetes Association (ADA) criteria as having normoglycemia (A1C < 5.7%) or prediabetes (5.7% ≤ A1C < 6.4%) (2). Demographic variables including age, sex, and race were obtained from structured self-reported enrollment data. Race categories with fewer than 10 participants, as well as individuals indicating mixed race or “other”, were grouped into a single “Other/Unknown” category for analysis.

**Figure 1.**
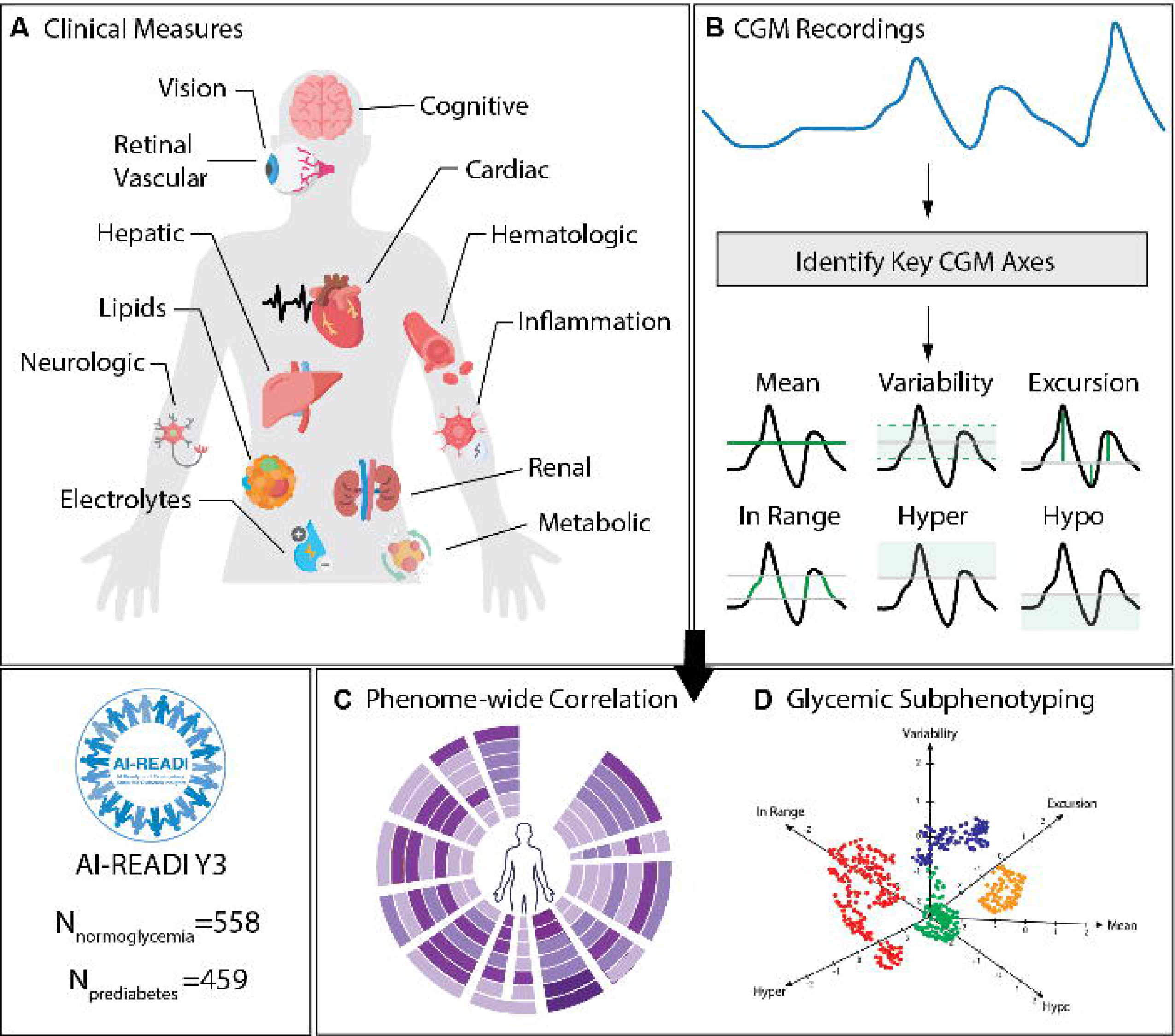
Study overview. (A) Clinical measures of end-organ alteration used for downstream analyses. (B) Continuous glucose monitoring (CGM) metrics were extracted from CGM recordings. Hierarchical clustering identified six glycemic axes, from which one representative metric per axis was selected for downstream analyses. (C) Phenome-wide pairwise correlation analysis between clinical measures and CGM metrics. (D) Glycemic subphenotyping based on CGM metrics.

### CGM Data Preprocessing

CGM data were processed by first excluding the initial 12 hours of wear to account for sensor equilibration. We removed partial recording days, defined as the first or last calendar days with fewer than 288 observations. Participants were included in the final analysis only if they maintained at least seven full days of data with a minimum of 70% daily coverage (at least 202 readings per 24-hour period). Out-of-range “High” and “Low” sensor readings were standardized to 401 mg/dL and 39 mg/dL, respectively, and participants with temporal gaps exceeding 12 consecutive hours were excluded. After preprocessing of the CGM readings, 52 CGM-derived metrics were extracted using the *iglu* package (19). To reduce redundancy among highly correlated metrics, a Spearman correlation matrix was constructed, and hierarchical clustering was performed using average linkage with Euclidean distance on the correlation structure (Supplementary Fig 1). Metrics were organized by cutting the hierarchical clustering dendrogram to yield six groups that represent different dimensions of glucose dynamics (Fig 1B). One representative measure was selected for each dimension following approaches used in prior literature (13). When available, metrics included in international consensus recommendations were prioritized (7). If multiple consensus metrics were present within a cluster, the measure with the narrower glycemic range was selected (e.g., time in tight range over time in range), as these measures are more sensitive to subtle glycemic variation in a non-diabetic population.

### Clinical Data Preprocessing

A total of 46 clinical measurements were obtained during the on-site assessment, including laboratory testing (blood and urine), physical examination, and monofilament testing. An additional 4 clinically relevant indices were derived: the triglyceride-to-high-density lipoprotein cholesterol ratio (TG/HDL) (20), triglyceride-glucose (TyG) index (21), urine albumin-to-creatinine ratio (UACR) (22), and estimated glomerular filtration rate (eGFR) calculated using the 2021 CKD-EPI creatinine equation (23). A total of 8 ECG waveform measures extracted by the Philips PageWriter TC30 software were included from the ECG assessment. For vision tests, measurements from the left (OS) and right (OD) eyes were averaged to generate 13 variables. Cognitive performance was assessed using the Montreal Cognitive Assessment (MoCA) (24), from which item-level scores and task completion times were available. To facilitate interpretability, MoCA components were aggregated into domain-specific measures, including visuospatial/executive function (trail making, cube copy, and clock drawing scores), attention/working memory (digit span, letter tapping, and serial subtraction scores), and language (naming, sentence repetition, and verbal fluency scores), while memory was represented using the combined Memory Index Score and orientation was retained as a separate domain. A processing speed measure was calculated as the sum of completion times for tasks with meaningful and consistently recorded response latencies, excluding fixed-duration or examiner-paced tasks, and cognitive efficiency was computed as the ratio of total MoCA score to processing speed. These transformations yielded 8 cognitive measures for subsequent analyses. Lastly, 7 measures were extracted from optical coherence tomography angiography (OCTA) imaging. In total, the analytical dataset comprised 86 measures organized into 13 end-organ domains. For selected analyses, a subset of was used to avoid redundancy among highly correlated measures or when variables served solely as intermediate components of derived indices.

### OCTA Data Preprocessing

We extracted measures of retinal vasculature from the superficial vascular complex (SVC) and deep vascular complex (DVC) from OCTA images. Because OCTA images are highly susceptible to motion, projection, and signal reduction artifacts, all scans were first reviewed by a clinician for quality control and exclusion assessment (25). To extract vasculature information, an nnU-Net pretrained on synthetic OCTA (26) images was applied for segmentation, followed by a graph extraction pipeline using described in our prior study (25). Different summary statistics were used to characterize the distribution of these metrics, yielding features such as total vessel length and the number of branching points. FAZ-related metrics were extracted from the segmented images, including FAZ area, perimeter, circularity, and acircularity index.

### Statistical Analysis

Baseline demographics, CGM metrics, and clinical measures were compared between participants with A1C-defined normoglycemia and prediabetes. Continuous variables were assessed for skewness (Fisher-Pearson coefficient >2.5 indicating right skew). Normally distributed variables were analyzed using Welch’s t-test and reported as mean ± SD with 95% CIs and Cohen’s d. Right-skewed positive variables were log-transformed and reported as geometric mean ratios with 95% CIs, whereas skewed variables containing non-positive values were analyzed using the Mann-Whitney U test and reported as median (IQR) with bootstrap-derived 95% CIs. Categorical variables were compared using Fisher’s exact or chi-square tests, as appropriate. Multiple comparisons were controlled using the Benjamini-Hochberg false discovery rate (FDR) procedure (p < 0.05).

### Phenome-wide Association Analysis

The first stage of the phenome-wide analysis evaluated associations between dysglycemic patterns and end-organ alterations by calculating partial Spearman correlation coefficients between each CGM metric and clinical measurement across all participants (Fig. 1C). Analyses were adjusted for age, BMI, and sex. Spearman correlation was chosen for its robustness to non-normality and outliers while preserving monotonic relationships. Correlation coefficients and corresponding p-values were computed for each CGM-clinical variable pair, with multiple comparisons controlled using FDR. Statistical significance was defined as adjusted p < 0.05.

Glycemic subphenotyping was then performed using standardized CGM metrics as input (Fig. 1D). Dimensionality reduction was conducted using UMAP (50 neighbors, minimum distance = 0.1, Canberra distance), followed by HDBSCAN clustering on the two-dimensional embedding (minimum cluster size = 60, minimum samples = 30) (27,28). Cluster quality was evaluated using manifold trustworthiness, cluster persistence, noise proportion, and stability across random initializations quantified by pairwise Adjusted Rand Indices (ARI). A column-shuffle null analysis preserving marginal feature distributions was performed to confirm that observed clustering exceeded chance expectations. The final solution demonstrated high neighborhood preservation (trustworthiness = 0.985), high stability (mean ARI = 0.940), and a low proportion of noise points (1.08%). Individuals were assigned to clusters based on maximum membership probability, with core members defined by posterior probability ≥0.7.

Differences in demographic and clinical variables across glycemic subphenotypes were evaluated using chi-square tests for categorical variables, one-way ANOVA for normally distributed variables, ANOVA on log-transformed values for strictly positive skewed variables, and Kruskal-Wallis tests for skewed variables with non-positive values. For significant chi-square tests, adjusted standardized residuals were used to identify over– and underrepresented categories (29). Effect sizes were reported as eta-squared for ANOVA and epsilon-squared for Kruskal-Wallis tests. Variables with significant omnibus results after FDR correction underwent pairwise testing using Tukey’s honestly significant difference test for parametric analyses or Mann-Whitney U tests with Benjamini-Hochberg adjustment for nonparametric analyses.

## Results

### Cohort Demographics and Baseline Characteristics

The analytic cohort included 1,017 participants, comprising 558 with normoglycemia and 459 with prediabetes defined by A1C thresholds (Table 1). Prediabetes was more prevalent among individuals aged 55–64 years, whereas sex distributions were similar between groups (61.1% vs. 62.9% female). Compared with the normoglycemic group, participants with prediabetes included lower proportions of White individuals (39.4% vs. 53.8%) and higher proportions of Black (19.7% vs. 12.3%) and Asian (35.4% vs. 25.7%) individuals. Baseline comparisons identified 34 significant differences in clinical measures (Table 1; Fig 1A) and 47 significant differences in CGM metrics (Supplementary Table 2; Fig 1B) after FDR correction. Nearly all CGM metrics (47/52, 90%) differed significantly, with only measures of extended hypoglycemia and temporal instability showing no significant differences.

**Table 1.** Baseline Demographic, Clinical, and CGM Characteristics by Glycemic Status.

| Variable | Normoglycemia | Prediabetes | Effect (95% CI) | P-value |
| --- | --- | --- | --- | --- |
| *n* | 558 | 459 |  |  |
| **Age (years)** |  |  |  | <0.001 |
| 35–44 | 79 (14.2%) | 26 (5.7%) |  |  |
| 45–54 | 159 (28.5%) | 99 (21.6%) |  |  |
| 55–64 | 124 (22.2%) | 145 (31.6%) |  |  |
| 65–74 | 146 (26.2%) | 138 (30.1%) |  |  |
| 75+ | 50 (9.0%) | 51 (11.1%) |  |  |
| **Sex** |  |  |  | 0.006 |
| Female | 350 (62.7%) | 280 (61.0%) |  |  |
| Male | 208 (37.3%) | 179 (39.0%) |  |  |
| **Race** |  |  |  | <0.001 |
| White | 301 (53.9%) | 182 (39.7%) |  |  |
| Asian | 144 (25.8%) | 163 (35.5%) |  |  |
| Black | 68 (12.2%) | 89 (19.4%) |  |  |
| **Other/Unknown** | 45 (8.1%) | 25 (5.4%) |  |  |
| **Ethnicity** |  |  |  | 0.13 |
| Hispanic | 24 (4.3%) | 8 (1.7%) |  |  |
| Non-Hispanic | 534 (95.7%) | 451 (98.3%) |  |  |
| **CGM Metrics** |  |  |  |  |
| TITR (%) | 87.42 (11.91) | 78.28 (18.68) | -9.14 (-11.11, -7.16) | <0.001 |
| Mean Glucose (mg/dL) | 115.71 (12.04) | 123.70 (15.79) | 7.99 (6.23, 9.75) | <0.001 |
| TAR‡ (%) | 0.52 [0.00, 1.55] | 1.56 [0.39, 4.88] | 1.04 (0.74, 1.34) | <0.001 |
| CV (%) | 16.32 (3.81) | 17.75 (4.19) | 1.44 (0.94, 1.93) | <0.001 |
| MAGE (mg/dL) | 48.70 (13.92) | 57.57 (18.29) | 8.87 (6.83, 10.90) | <0.001 |
| LBGI‡ | 0.32 [0.14, 0.72] | 0.19 [0.06, 0.52] | -0.13 (-0.19, -0.08) | <0.001 |
| **Body Composition** |  |  |  |  |
| Waist Circumference (cm) | 94.66 (15.47) | 97.62 (17.45) | 2.96 (0.91, 5.01) | <0.01 |
| Waist-to-Height Ratio | 0.89 (0.09) | 0.91 (0.11) | 0.02 (0.00, 0.03) | <0.05 |
| **Metabolic** |  |  |  |  |
| HbA1c (%) | 5.36 (0.22) | 5.90 (0.20) | 0.54 (0.51, 0.56) | <0.001 |
| Glucose (mg/dL) | 88.67 (15.97) | 96.82 (16.92) | 8.15 (6.10, 10.19) | <0.001 |
| C-Peptide (ng/mL) | 2.64 (1.86) | 3.03 (2.11) | 0.39 (0.14, 0.64) | <0.01 |
| Insulin† (ng/mL) | 0.47 | 0.59 | 1.26 (1.12, 1.41) | <0.001 |
| Triglyceride/HDL† | 1.90 | 2.22 | 1.17 (1.08, 1.27) | <0.001 |
| Triglyceride-Glucose Index | 8.50 (0.55) | 8.70 (0.57) | 0.20 (0.13, 0.26) | <0.001 |
| **Lipids** |  |  |  |  |
| Triglycerides (mg/dL) | 128.25 (73.23) | 143.34 (78.18) | 15.09 (5.70, 24.49) | <0.01 |
| HDL Cholesterol (mg/dL) | 61.44 (16.33) | 58.50 (15.37) | -2.94 (-4.90, -0.99) | <0.01 |
| **Renal** |  |  |  |  |
| BUN† (mg/dL) | 14.56 | 15.55 | 1.07 (1.03, 1.11) | <0.001 |
| Urine Albumin‡ (mg/mL) | 0.33 [0.08, 0.73] | 0.40 [0.15, 0.90] | 0.07 (0.01, 0.15) | <0.01 |
| **Hepatic** |  |  |  |  |
| Alkaline Phosphatase (IU/L) | 69.47 (20.54) | 72.88 (22.04) | 3.41 (0.77, 6.06) | <0.05 |
| AST† (IU/L) | 19.33 | 20.34 | 1.05 (1.01, 1.09) | <0.01 |
| ALT† (IU/L) | 17.95 | 19.95 | 1.11 (1.05, 1.18) | <0.001 |
| Protein, total (g/dL) | 6.92 (0.44) | 7.01 (0.44) | 0.09 (0.04, 0.15) | <0.01 |
| Globulin (g/dL) | 2.56 (0.38) | 2.67 (0.39) | 0.11 (0.06, 0.16) | <0.001 |
| A/G Ratio | 1.74 (0.29) | 1.66 (0.27) | -0.08 (-0.11, -0.04) | <0.001 |
| **Hematologic** |  |  |  |  |
| MCV (fL) | 91.44 (5.39) | 90.08 (5.48) | -1.36 (-2.03, -0.69) | <0.001 |
| MCH (pg) | 30.26 (2.17) | 29.57 (2.21) | -0.69 (-0.96, -0.42) | <0.001 |
| MCHC (g/dL) | 33.09 (1.06) | 32.81 (1.01) | -0.28 (-0.40, -0.15) | <0.001 |
| Red Cell Distribution Width (%)† | 13.00 | 13.31 | 1.02 (1.01, 1.03) | <0.001 |
| **Vision** |  |  |  |  |
| Photopic Letter Score | 84.29 (7.18) | 83.11 (7.40) | -1.18 (-2.08, -0.28) | <0.05 |
| Photopic LogMAR | 0.01 (0.14) | 0.04 (0.15) | 0.02 (0.01, 0.04) | <0.05 |
| Mesopic Letter Score | 72.49 (7.92) | 71.10 (8.00) | -1.39 (-2.38, -0.41) | <0.01 |
| Mesopic LogMAR | 0.25 (0.16) | 0.28 (0.16) | 0.03 (0.01, 0.05) | <0.01 |
| Mesopic Final Letter Value | 1.35 (0.17) | 1.31 (0.19) | -0.04 (-0.06, -0.02) | <0.001 |
| Mesopic Log Contrast Sensitivity | 1.32 (0.17) | 1.28 (0.18) | -0.04 (-0.06, -0.01) | <0.01 |
| **Cognitive (MoCA)** |  |  |  |  |
| Total Score | 26.48 (2.70) | 25.76 (2.95) | -0.72 (-1.08, -0.37) | <0.001 |
| Processing Speed | 3.57 (0.30) | 3.67 (0.30) | 0.10 (0.06, 0.14) | <0.001 |
| Efficiency | 0.73 (0.27) | 0.64 (0.24) | -0.09 (-0.13, -0.06) | <0.001 |
| Attention | 5.55 (0.87) | 5.39 (0.98) | -0.16 (-0.27, -0.04) | <0.01 |
| Language | 5.03 (1.04) | 4.79 (1.10) | -0.25 (-0.38, -0.11) | <0.001 |
| Memory | 13.01 (2.51) | 12.58 (2.78) | -0.43 (-0.76, -0.10) | <0.05 |

† Reported as geometric mean and geometric mean ratio with 95% CI.
‡ Reported as median [IQR]; effect represents median difference with 95% CI.

As expected, individuals with prediabetes had greater baseline central adiposity, reflected by higher waist circumference (97.6 vs. 94.7 cm, p<0.01), along with a more atherogenic and insulin-resistant profile characterized by higher triglycerides (143 vs. 128 mg/dL, p<0.01) and lower HDL cholesterol (58.5 vs. 61.4 mg/dL, p<0.01) (Table 1). Notably, no significant difference in mean BMI was observed (28.9 vs. 28.0, p=0.10).

Individuals with prediabetes exhibited modest but significant differences across several clinical measures, including higher BUN (15.55 vs. 14.56 mg/dL, p<0.001), ALT (19.95 vs. 17.95 IU/L, p<0.001), and Red Cell Distribution Width (RDW) (13.3% vs. 13.0%, p<0.001). They also had a lower total Montreal Cognitive Assessment (MoCA) score (25.8 vs. 26.5, p<0.001) and higher photopic LogMAR values (0.04 vs. 0.01, p<0.05), indicating poorer cognitive and visual function. Complete baseline comparisons of all 86 clinical measures are provided in Supplementary Table 1.

Because many CGM metrics are highly related in mathematical definition, we performed hierarchical clustering on the pairwise Spearman correlation matrix of 52 metrics and identified six nonredundant axes of glycemic control (Fig 1B). One representative metric from each axis was selected for downstream analyses (Supplementary Fig 1, green highlights): Mean Glucose, Coefficient of Variation (CV), Mean Amplitude of Glycemic Excursions (MAGE), Time in Tight Range (TITR; 63–140 mg/dL), Time Above Range >180 mg/dL (TAR), and Low Blood Glucose Index (LBGI). Although selected to reduce redundancy, these representative metrics can capture complementary aspects of glycemic physiology, including hyperglycemia (Mean Glucose, TITR, TAR), glycemic variability (CV, MAGE), and hypoglycemia risk (LBGI).

### End-Organ Alterations in Early Dysglycemia

We assessed 462 correlations between 13 categories of clinical measures and 7 glycemic metrics, of which 99 (21.4%) were significant after FDR correction. The strongest associations were observed for cardiometabolic, cardiovascular, and cognitive measures (Fig 2A). Overall, metrics reflecting elevated glycemia, including Mean Glucose, TAR, and TITR, demonstrated substantially more significant associations than measures of glycemic variability, particularly CV (Fig 2B). Consistent with prior evidence linking insulin resistance to hyperglycemia and glucose excursions (30), C-peptide correlated with TITR (ρ = 0.15, p<0.001), Mean Glucose (ρ = 0.12, p<0.001), and MAGE (ρ = 0.12, p<0.001), but not CV, a pattern also observed for triglycerides and C-reactive protein (CRP).

**Figure 2.**
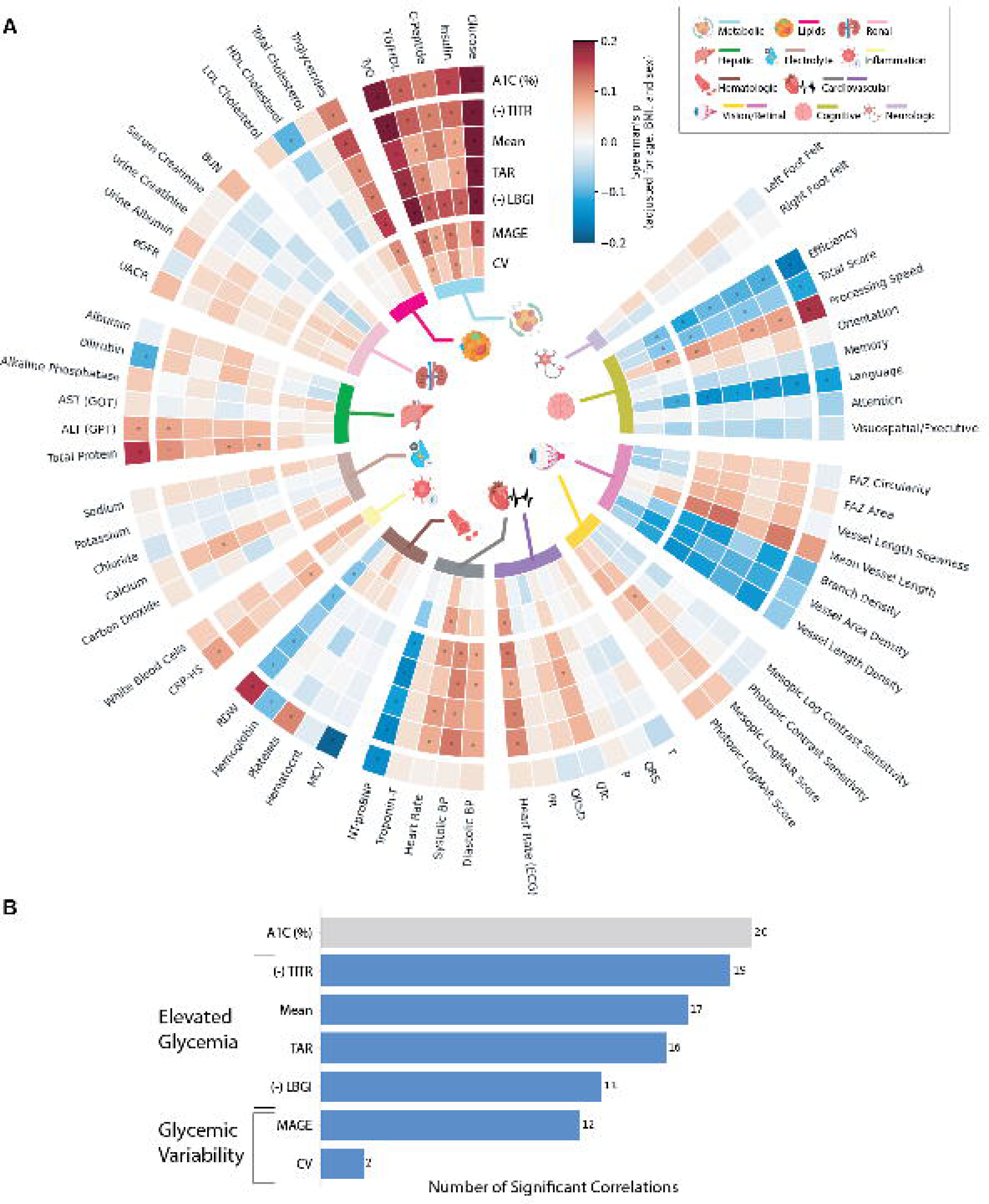
Phenome-wide correlations between A1C and glycemic metrics. (A) Circular heatmap showing correlation strength between A1C, CGM metrics, and clinical measures spanning 13 end-organ systems. Rows represent glycemic metrics, including HbA1c, time in tight range (TITR), mean glucose, time above range (TAR), low blood glucose index (LBGI), mean amplitude of glycemic excursions (MAGE), and coefficient of variation (CV). Columns represent individual clinical measures grouped by end-organ system, as indicated by the colored inner ring. Associations were quantified using Spearman rank correlation coefficients adjusted for age, sex, BMI, and race among 1,017 participants. Color intensity reflects the direction and magnitude of association (blue, negative; red, positive), and dots denote associations that remained significant after false discovery rate (FDR) correction. For visualization, TITR and LBGI were sign-flipped such that higher values consistently indicate poorer glycemic profiles. (B) Number of significant associations identified for each glycemic metric after FDR correction.

Renal and hepatic dysfunction are recognized complications of diabetes and may emerge during early dysglycemia (31,32). Consistent with this, individuals with prediabetes exhibited significant baseline differences in several renal and hepatic markers (Table 2). However, few markers were associated with CGM metrics (Fig 2A): no renal markers showed significant correlations, and among hepatic markers, only ALT was modestly associated with Mean Glucose (ρ = 0.09, p < 0.05). This suggests that early renal and hepatic abnormalities may reflect cumulative hyperglycemic exposure rather than short-term glycemic patterns captured by CGM.

In contrast, several cardiovascular indices were associated with CGM metrics despite showing no significant differences between groups. Systolic and diastolic blood pressure were positively correlated with Mean and TITR (all p<0.05), while NT-proBNP was inversely correlated with Mean Glucose (ρ = −0.15, p<0.001) and TAR (ρ = −0.12, p<0.001). Among ECG measures, we identified a significant correlation between QTc and LBGI (ρ = 0.08, p<0.05) and moderate but non-significant correlations with Mean Glucose and TAR (Fig 2A). Although prior studies on QTc prolongation in prediabetes have reported mixed findings (33), our results align with the hypothesis that hyperglycemia may impair ventricular repolarization through prolongation of sarcolemma action potentials (34).

Additionally, both ECG-derived and in-clinic heart rate measurements were significantly correlated with Mean Glucose, TAR, and TITR, consistent with prior evidence linking increasing A1C to elevated heart rate and altered autonomic regulation (35). Notably, none of the cardiovascular measures were associated with A1C, suggesting greater sensitivity to transient hyperglycemia captured by CGM than to longer-term glycemic exposure.

RDW differed significantly between normoglycemic and prediabetic participants and was correlated with both CGM metrics and A1C. However, the direction of association differed: RDW was inversely correlated with Mean Glucose, TAR, MAGE, and TITR (all ρ = −0.09, p < 0.05) but positively correlated with A1C (ρ = 0.16, p < 0.001). These opposing relationships suggest that RDW may be influenced by long-term glycation and erythrocyte turnover, consistent with prior studies linking RDW to future A1C elevation rather than glucose elevation (36).

A unique aspect of the AI-READI dataset is the inclusion of visual and cognitive assessments. Cognitive impairment in diabetes, particularly affecting attention, memory, and executive function, has been linked to impaired insulin signaling, white matter changes, and subclinical vascular injury (37). However, these relationships remain understudied in early dysglycemia. Mesopic LogMAR was correlated with TITR (ρ = 0.09, p < 0.05), while total MoCA score, language score, processing speed, and efficiency were correlated with TITR, MAGE, and TAR (ρ ≈ −0.10 to −0.14, p < 0.01). These findings suggest that sustained hyperglycemia and glycemic excursions are associated with poorer low-light visual acuity and cognitive performance. Notably, language function was the only cognitive subdomain associated with both CGM metrics and A1C, with stronger associations observed for CGM metrics. This finding is particularly noteworthy given the limited and inconsistent literature on language impairment compared with the more extensively studied executive function and memory domains (37).

### Data-Driven Glycemic Subphenotyping

To determine whether CGM-derived glycemic axes define reproducible subphenotypes with potential utility for risk stratification, we applied unsupervised machine learning to the representative CGM metrics and identified four distinct subphenotypes: Healthy (N=154, Cluster Persistence (CP)=0.344), Mild Hyperglycemia (N=155, CP=0.406), High Variability (N=199, CP=0.452), and Hyperglycemia (N=475, CP=0.483) (Fig 3A). The Hyperglycemia phenotype exhibited persistently elevated glycemia, with the highest Mean Glucose, MAGE, and TAR and the lowest TITR (Fig 3C, red), whereas the High Variability phenotype was characterized by the highest CV despite lower Mean Glucose and TAR (Fig 3C, orange). Healthy demonstrated the most favorable glycemic profile, with high TITR and low Mean Glucose, CV, and MAGE, while Mild Hyperglycemia occupied an intermediate position with modest elevations in Mean Glucose and TAR but the lowest CV overall (Fig 3C, green/blue). Clinical characteristics of the subphenotypes are summarized in Supplementary Table 4.

**Figure 3.**
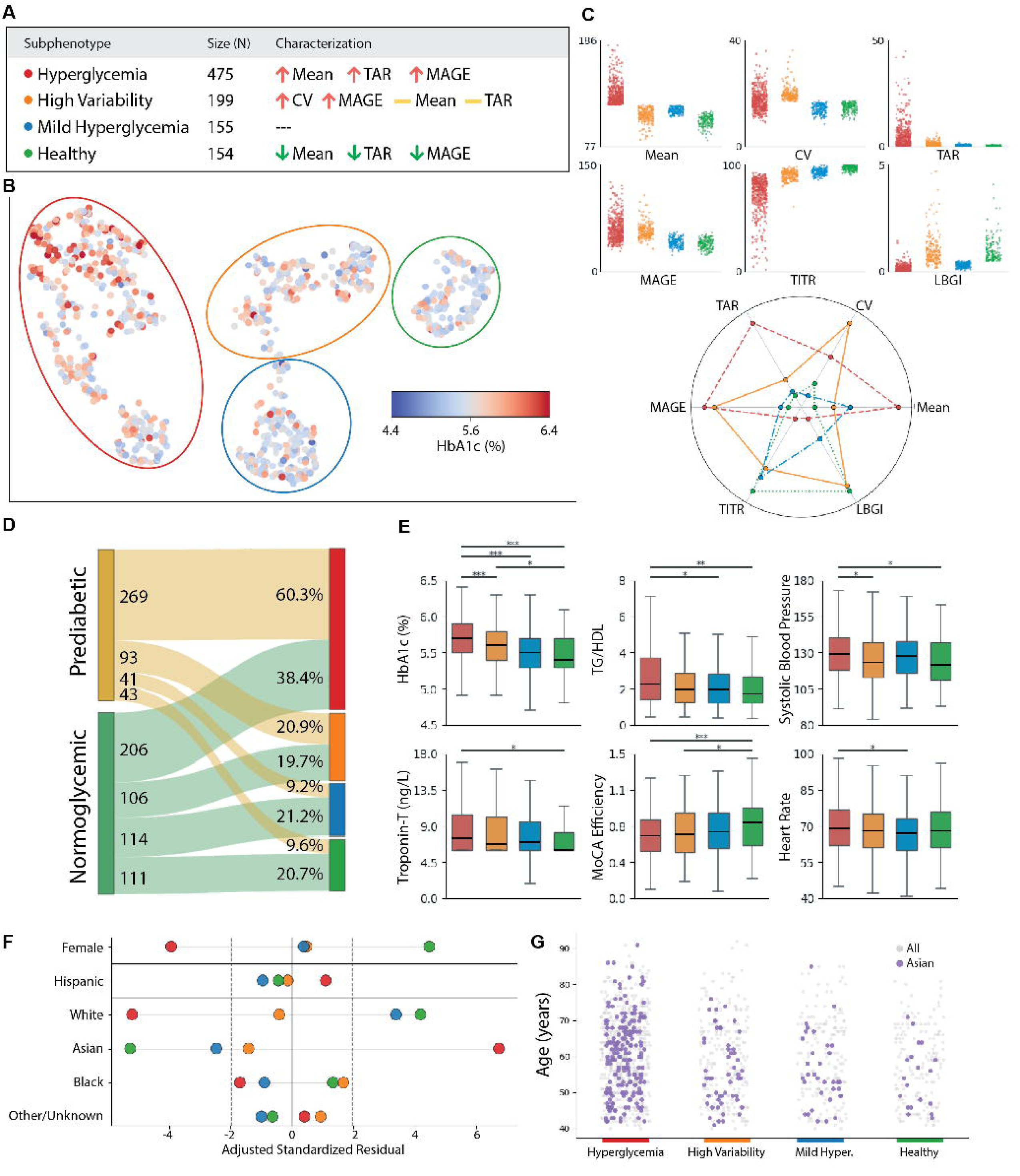
Identification and characterization of glycemic subphenotypes. (A) Summary of the four CGM-derived glycemic subphenotypes and their defining glycemic characteristics. (B) UMAP embedding of participants based on CGM metrics. Each point represents an individual participant and is colored by A1C, with lower values shown in blue and higher values shown in red. Colored outlines indicate the four clusters identified by HDBSCAN. (C) Distribution and radar plots of CGM metrics across subphenotypes, including mean glucose, coefficient of variation (CV), time above range >180 mg/dL (TAR), mean amplitude of glycemic excursions (MAGE), time in tight range (TITR), and low blood glucose index (LBGI). (D) Sankey diagram showing the relationship between A1C-defined glycemic status and glycemic subphenotypes. (E) Comparison of selected clinical and functional measures across subphenotypes. Asterisks denote statistical significance (*P < 0.05, **P < 0.01, ***P < 0.001). (F) Adjusted standardized residuals showing over– and underrepresentation of demographic groups across subphenotypes. Vertical dashed lines indicate the significance threshold (adjusted standardized residual = ±1.96). (G) Age distributions across glycemic subphenotypes. Distributions for Asian participants (purple) are overlaid on overall cluster distributions (gray).

To compare CGM-derived subphenotypes with conventional A1C-based classification, we examined the distribution of A1C-defined normoglycemic and prediabetic individuals across the four glycemic subphenotypes (Fig 3D). Notably, 58.1% of normoglycemic individuals were classified into the dysglycemic Hyperglycemia (38.4%, N=206) or High Variability (19.7%, N=106) phenotypes. Conversely, despite elevated A1C levels, 18.8% (N=84) of prediabetic individuals were classified into the more favorable Healthy or Mild Hyperglycemia phenotypes. These findings highlight substantial heterogeneity within A1C-defined glycemic groups that is captured by CGM-based phenotyping.

Demographic composition differed significantly across glycemic subphenotypes by age, sex, and race (chi-square test, p < 0.001; Supplementary Table 3). The Healthy phenotype was younger and predominantly female (77.9% vs. 61.9% overall), with greater representation of individuals aged 35–44 years and lower representation of those aged ≥75 years. Adjusted standardized residual analysis revealed overrepresentation of Asian participants in the Hyperglycemia phenotype and White participants in the Healthy and Mild Hyperglycemia phenotypes (Fig 3F). The higher representation of Asian participants in the Hyperglycemia phenotype was consistent across age groups (Fig 3G).

Among the four subphenotypes, Hyperglycemia exhibited the most adverse cardiometabolic profile, with higher TG/HDL, TyG, systolic blood pressure, heart rate, and Troponin-T than the Healthy and Mild Hyperglycemia phenotypes (Fig 3E). These differences occurred despite no significant variations in body composition across subphenotypes (Supplementary Table 4). This suggests that CGM-defined hyperglycemia may identify individuals with elevated cardiometabolic risk not captured by body composition alone.

Cognitive differences also emerged across glycemic subphenotypes. Compared with Healthy, the Hyperglycemia phenotype exhibited lower total MoCA score, language score, and processing speed (Fig 3E), consistent with results from the phenome-wide correlation analysis (Fig 2). Notably, the High Variability phenotype also demonstrated lower MoCA efficiency and language scores despite a comparatively favorable metabolic profile. Because substantial glycemic variability can occur despite relatively normal A1C levels, these results suggest that cognitive risk may not be fully captured by conventional A1C-based classification.

## Conclusions

Early dysglycemia is associated with an increased risk of type 2 diabetes and end-organ alterations (12,13). Additionally, early dysglycemia may exhibit glycemic heterogeneity that contributes to differences in organ vulnerability. Here, we show that CGM identifies distinct axes of glycemic physiology associated with differential patterns of end-organ alteration, indicating that clinically relevant manifestations of dysglycemia emerge before overt diabetes and vary according to glycemic phenotype.

Across 13 end-organ categories, glycemic patterns were associated with a broad range of clinical measures. In addition to expected cardiometabolic and cardiovascular correlates, significant associations were observed in less-studied domains, including vision and cognition (6). Cognitive efficiency and language function declined with worsening dysglycemia, with language emerging as the cognitive domain most consistently associated with glycemic measures. This finding is notable given the mixed and inconsistent literature regarding language dysfunction in dysglycemia relative to attention, memory, and executive function (37). Because language assessments were conducted in English, these associations may be influenced by native language, educational attainment, and other sociodemographic factors. Nevertheless, cognitive alterations may represent an underrecognized manifestation of early dysglycemia and warrant further investigation.

Phenome-wide analysis revealed that distinct glycemic axes were differentially associated with end-organ alterations. Consistent with prior observations (13), both sustained and intermittent hyperglycemia emerged as the dominant correlates of end-organ alterations, suggesting that cumulative glucose exposure may be more closely linked to these alterations than glycemic variability. However, the clinical measures evaluated may incompletely capture processes associated with glycemic variability, such as endothelial dysfunction, oxidative stress, and inflammation, potentially underestimating its clinical relevance (30).

The apparent contribution of glycemic variability also depended on how variability was quantified. Although both MAGE and CV are widely used measures of variability, MAGE was associated with more clinical measures than CV. Because MAGE captures the magnitude of glucose excursions whereas CV reflects overall variability around the mean, these results suggest that excursion magnitude may be more relevant to early organ alterations than variability alone. While MAGE has previously been shown to distinguish normoglycemia from prediabetes (38), our findings suggest it may also provide insight into the clinical manifestations of dysglycemia. Future studies should further examine the direction, magnitude, and duration of glucose excursions (39), which may improve characterization and subphenotyping of early dysglycemia.

When examining the demographic distribution across the glycemic subphenotypes, we found that Asian individuals were overrepresented in the Hyperglycemia phenotype. This observation is consistent with prior evidence demonstrating a higher prevalence of prediabetes, insulin resistance, and type 2 diabetes among Asian populations, often occurring at lower BMI thresholds than in other racial and ethnic groups (40). This finding raises the possibility that CGM interpretation may benefit from population-specific approaches and supports further evaluation of CGM as a screening tool for identifying elevated glycemic risk despite normoglycemic A1C levels.

There is growing interest in using CGM to characterize aspects of glycemic physiology not captured by conventional measures such as A1C, particularly in early dysglycemia. Prior studies have linked CGM metrics to diabetes progression, vascular outcomes, and mortality, underscoring the clinical relevance of glycemic patterns beyond average glucose exposure (13). Yet, the extent to which these metrics reflect broader physiological alterations associated with early dysglycemia remains unclear. As CGM becomes increasingly accessible, addressing this gap will help define its role in the early detection and characterization of dysglycemia.

This study has several notable strengths. Using the AI-READI cohort, which comprises a large, demographically diverse population with balanced representation across A1C-defined normoglycemia and prediabetes, we were able to investigate glycemic heterogeneity across the early dysglycemia spectrum. First, we identified a set of glycemic axes that capture complementary dimensions of glycemic physiology while reducing redundancy among commonly reported CGM metrics. Second, we used these axes to perform a phenome-wide correlation analysis across a broad range of clinical measures, enabling a systematic assessment of how distinct glycemic patterns relate to end-organ alterations. This approach allowed us to examine CGM metrics not only as a tool for glycemic assessment, but also as a means of understanding the broader clinical manifestations of early dysglycemia. Third, we identified data-driven, CGM-derived glycemic subphenotypes with distinct clinical and demographic profiles, demonstrating that glycemic heterogeneity is associated with meaningful differences in clinical measures.

Several limitations should be acknowledged. The cross-sectional nature of the study precludes conclusions regarding causality or the temporal relationship between glycemic patterns and the observed end-organ alterations. In addition, CGM measurements were obtained during a single monitoring period, and the long-term stability of the identified glycemic phenotypes remains to be established. Furthermore, oral glucose tolerance testing was not available, precluding comparison of the identified glycemic subphenotypes with established prediabetes subtypes such as IGT and IFG. Future longitudinal studies will be particularly valuable for determining whether these CGM-derived measures and phenotypes predict progression to diabetes and the development of downstream complications. Finally, the clinical actionability of these findings remains to be defined – while identifying early signs of end-organ alterations is valuable, effective interventions specific to preventing such changes is an important next step.

## Supporting information

Supplementary Table 1 (continued)

Supplementary Table 4 (continued)

## Data Availability

All data produced are available online at

https://aireadi.org/dataset

## Acknowledgements

**Author Contributions and Guarantor Statement** B.C. conceived the study, conducted the analyses, and drafted the manuscript. W.T.L. and K.T. contributed to OCTA image analysis. J.D. and A.M. supervised the work and made substantial contributions to data interpretation and manuscript revision. All authors approved the final manuscript. B.C. and J.D. are the guarantors of this work and, as such, had full access to all study data and take responsibility for the integrity of the data and the accuracy of the analyses.

## Notes

### Competing Interest Statement

The authors have declared no competing interest.

### Author Declarations

The study uses only openly available human data that were originally located at: https://aireadi.org/dataset. Note: demographic data used in this study is a controlled portion of the AI-READI dataset which requires an additional approval process.

