## Supplementary figures and images for "Continuous Glucose Monitoring Reveals Glycemic Patterns Associated with End-Organ Alterations in Early Dysglycemia"

### Supplementary Figure 1

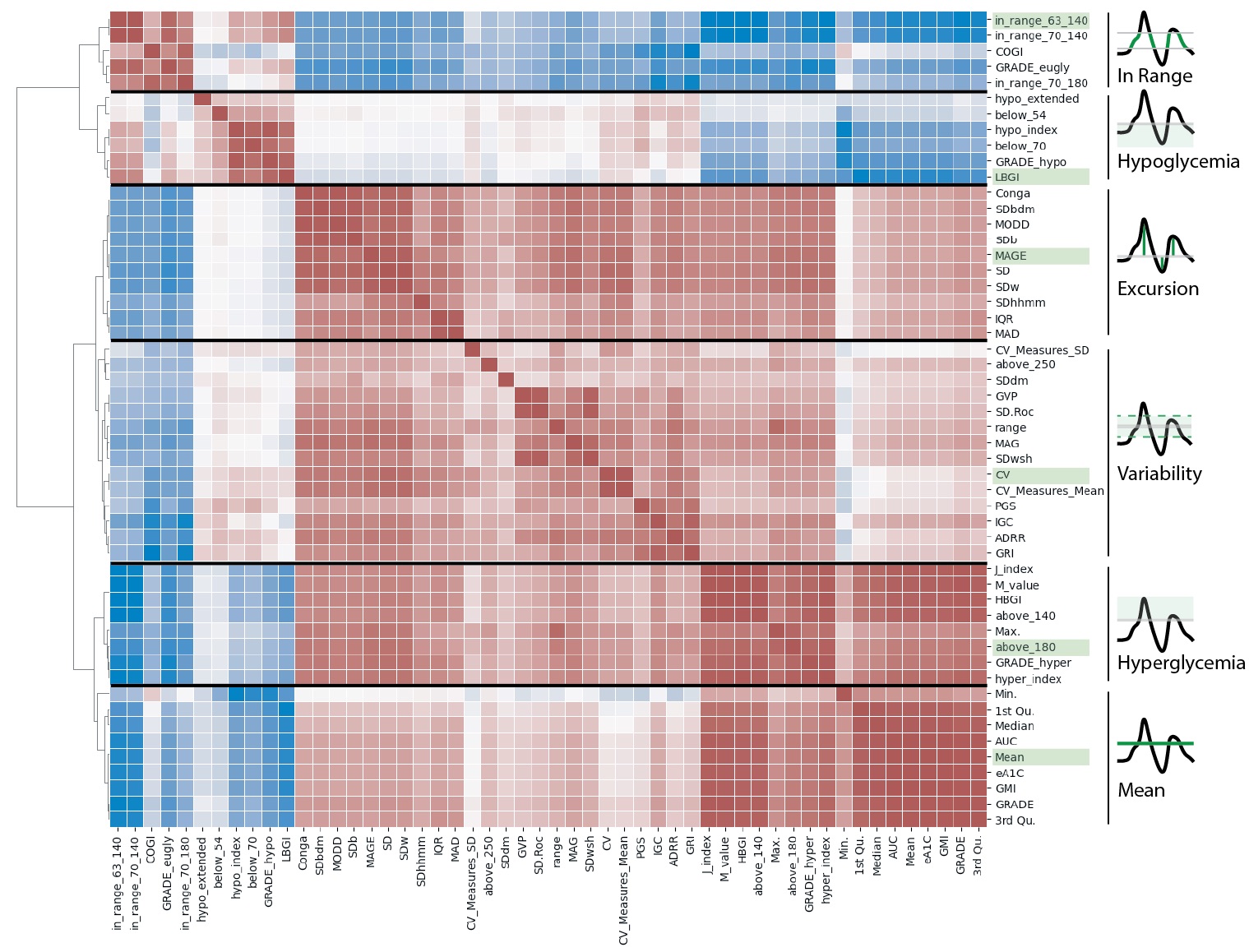

### Supplementary Table 1

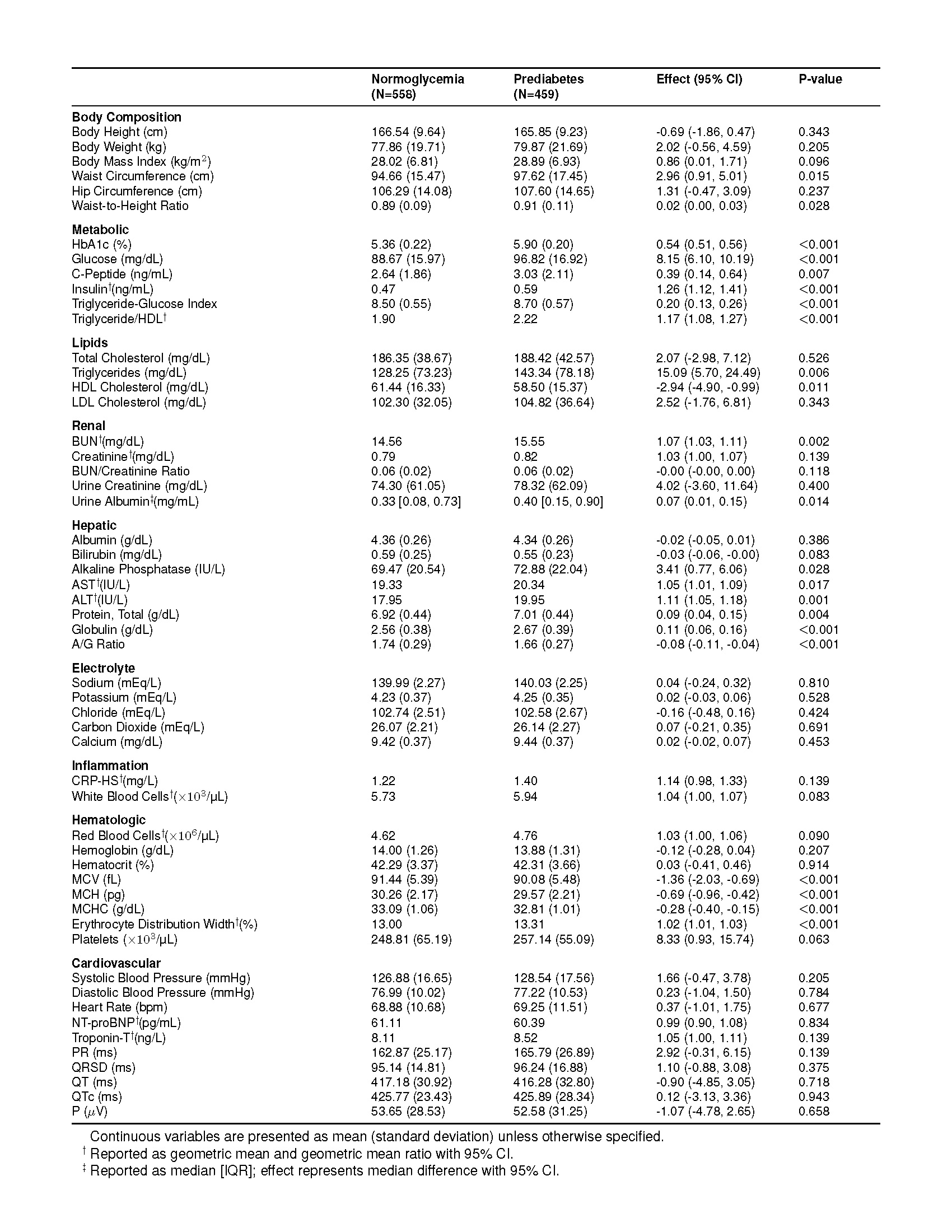

### Supplementary Table 1 (continued)

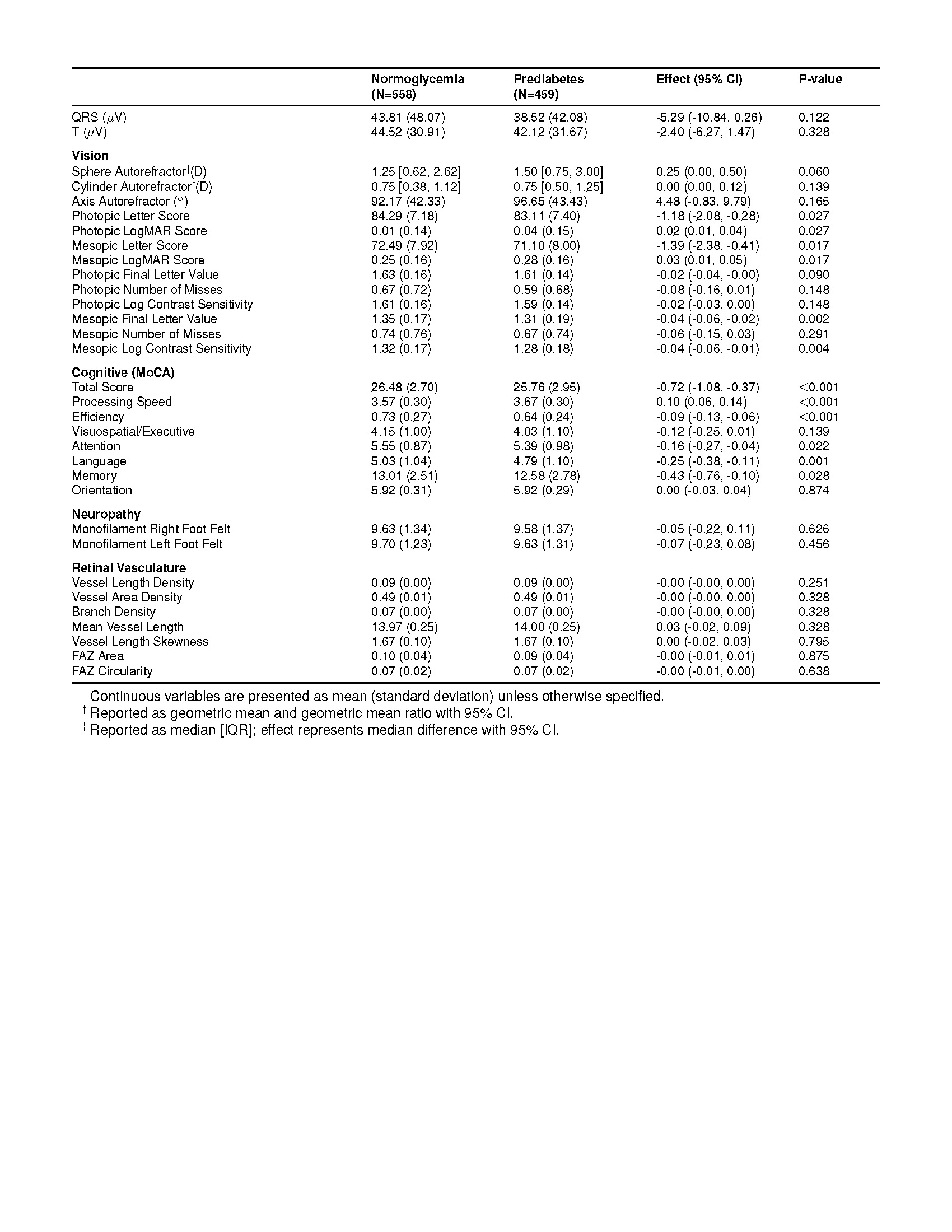

### Supplementary Table 2

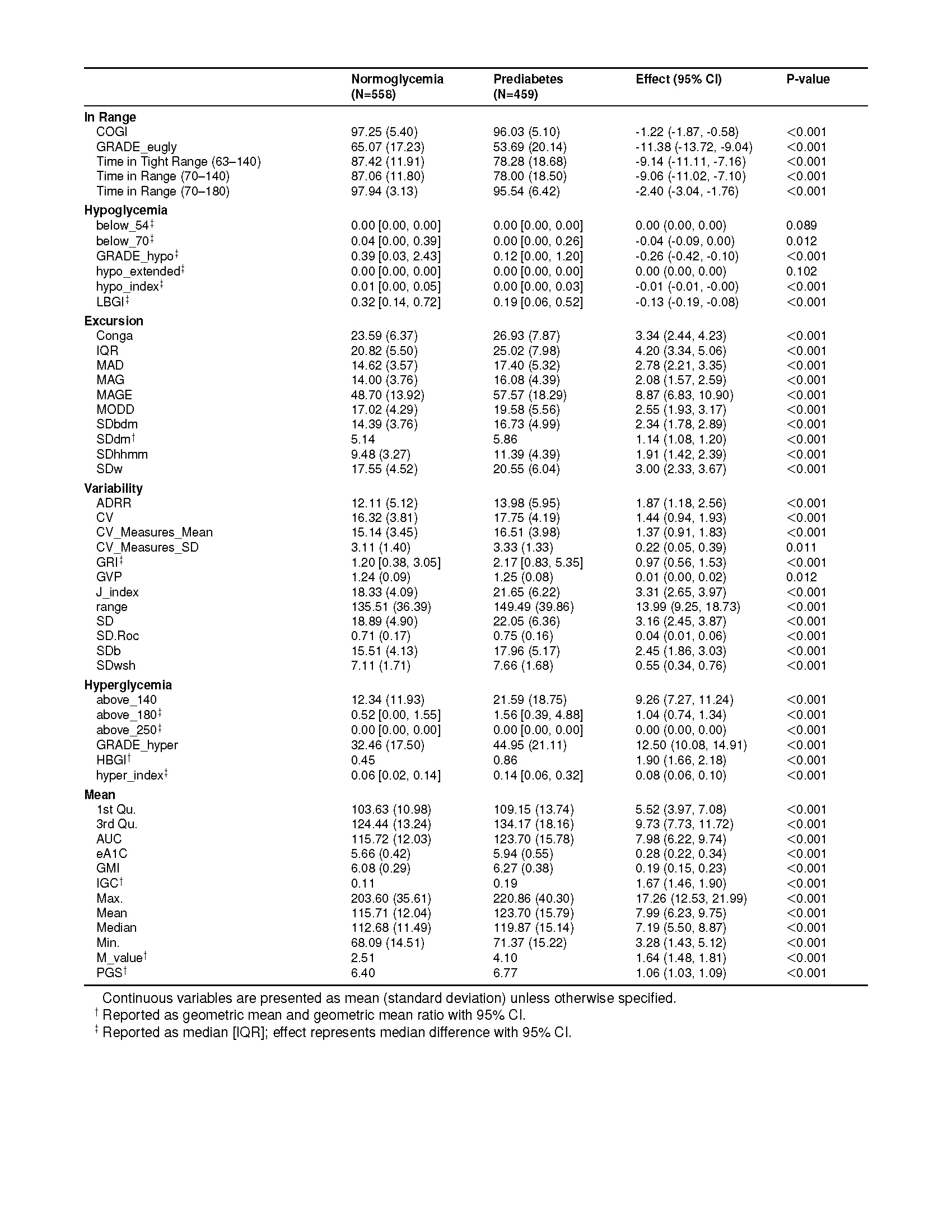

### Supplementary Table 3

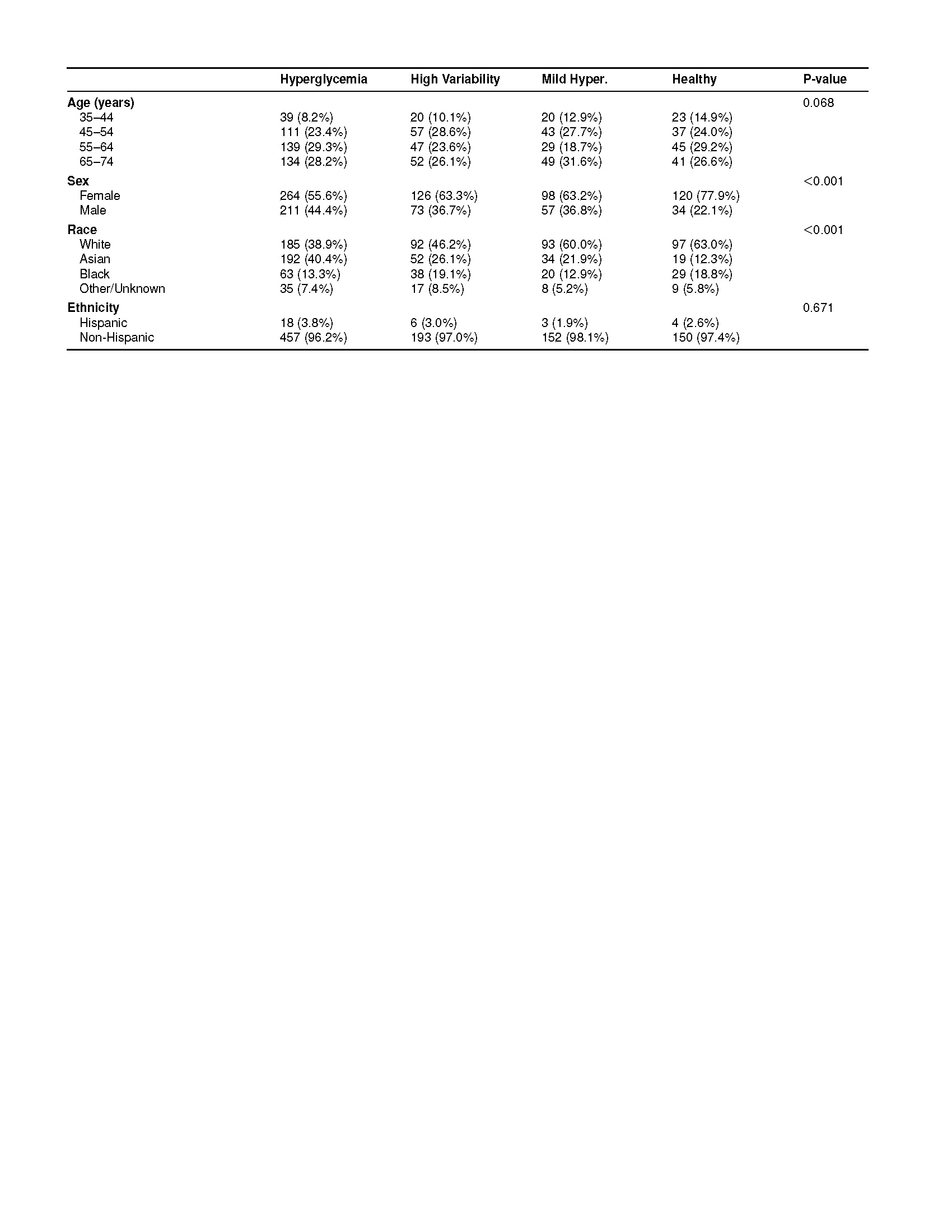

### Supplementary Table 4

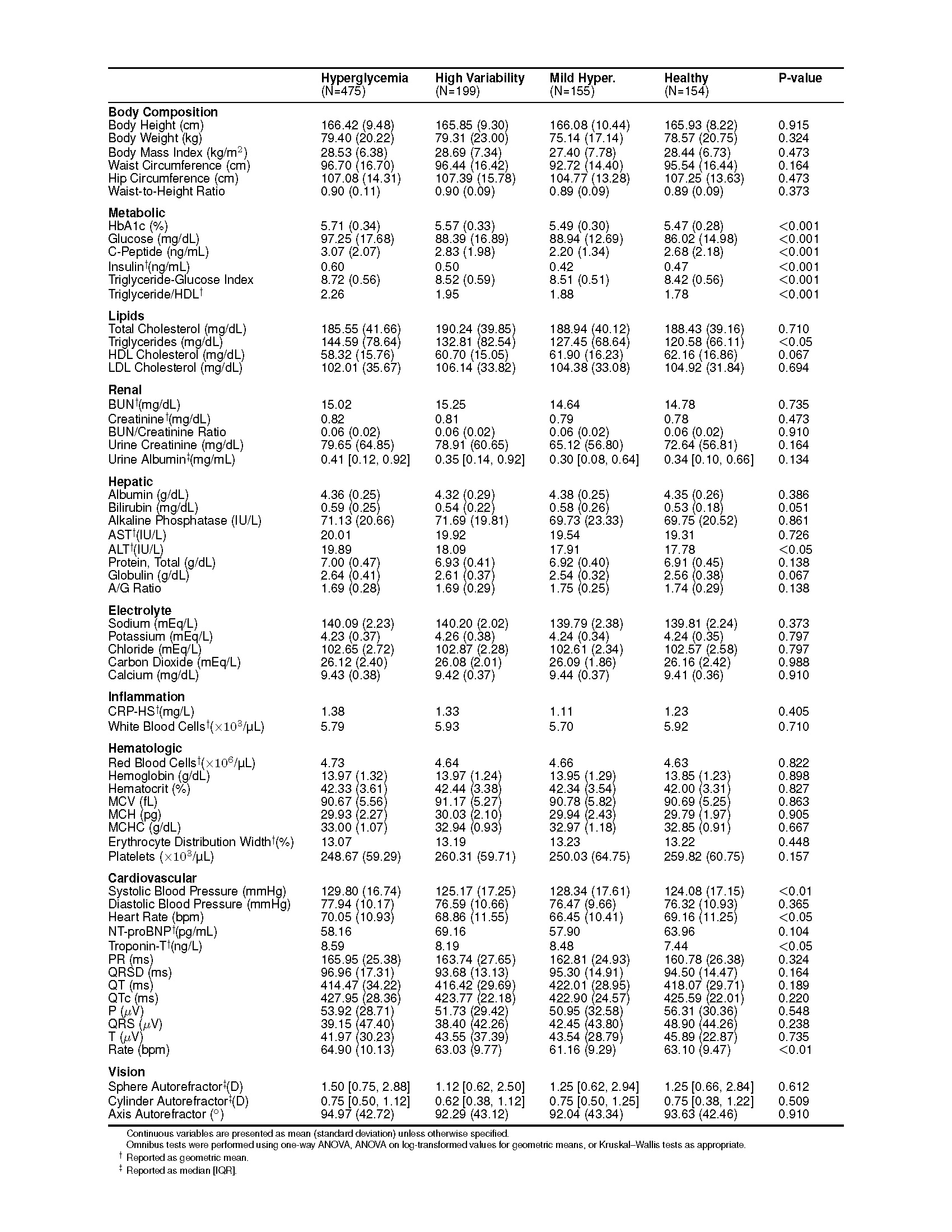

### Supplementary Table 4 (continued)

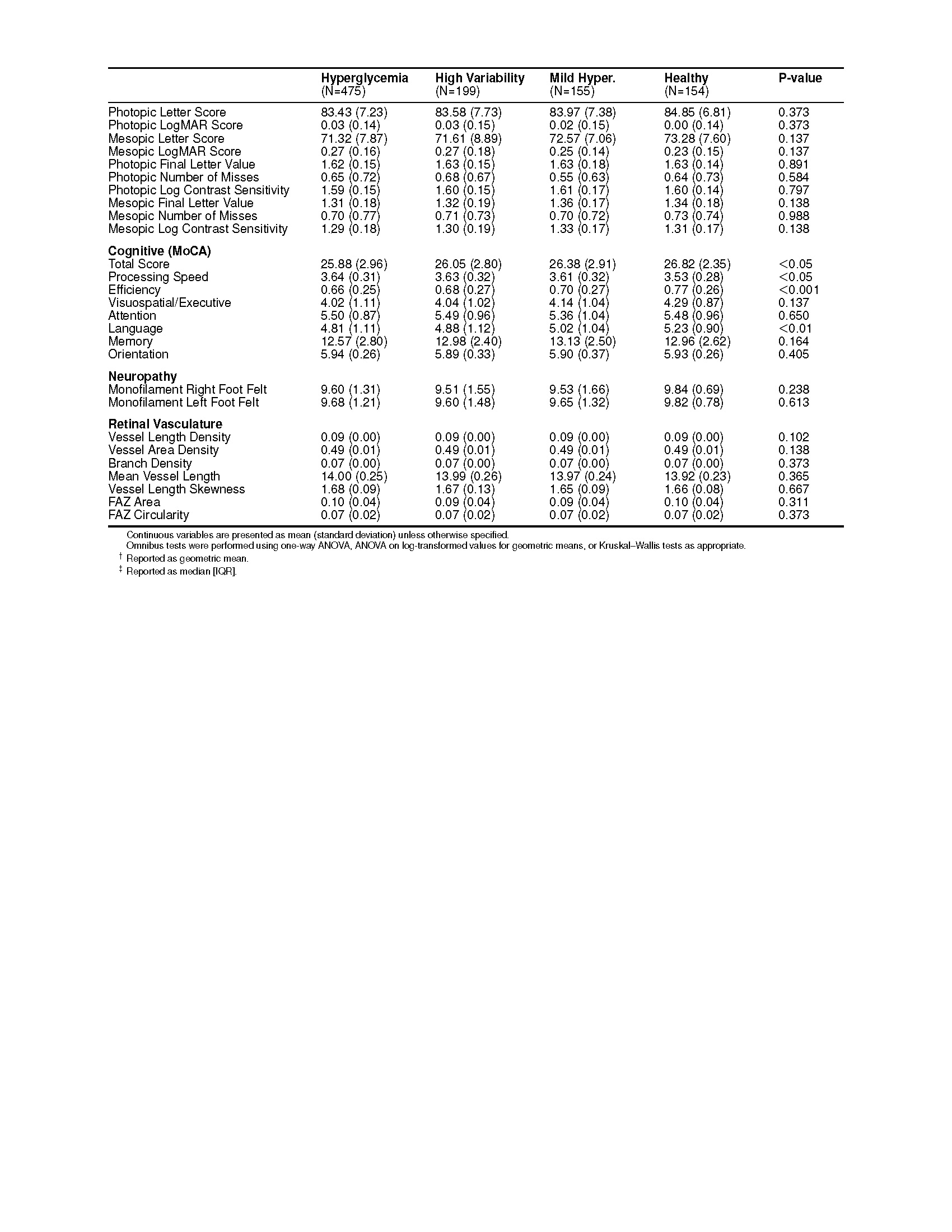
